# Linking air pollution exposure to blood-based metabolic features in a community-based aging cohort

**DOI:** 10.1101/2022.12.29.22284045

**Authors:** Vrinda Kalia, Erin R. Kulick, Badri Vardarajan, Yian Gu, Jennifer J. Manly, Mitchell S.V. Elkind, Joel D. Kaufman, Dean P. Jones, Andrea A. Baccarelli, Richard P. Mayeux, Marianthi-Anna Kioumourtzoglou, Gary W. Miller

**Affiliations:** Department of Environmental Health Sciences, Mailman School of Public Health, Columbia University. 722 West 168th Street, New York, NY 10032; Department of Epidemiology and Biostatistics, Temple University College of Public Health. 1301 Cecil B. Moore Avenue, Philadelphia, PA 19122; Taub Institute for Research on Alzheimer’s Disease and the Aging Brain, Vagelos College of Physicians and Surgeons, Columbia University. 630 West 168th Street, New York, NY 10032; The Gertrude H. Sergievsky Center, Vagelos College of Physicians and Surgeons, Columbia University. 630 West 168th Street, New York, NY 10032; Department of Neurology, Vagelos College of Physicians and Surgeons, Columbia University and the New York Presbyterian Hospital. 710 West 168th Street, New York, NY 10032; Department of Epidemiology, Mailman School of Public Health, Columbia University. 722 West 168th Street, New York, NY 10032; Departments of Environmental and Occupational Health Sciences, Medicine, and Epidemiology, University of Washington. 3980 15th Ave NE, Seattle, WA 98195; Clinical Biomarkers Laboratory, Department of Medicine, Emory University. 615 Michael Street, Atlanta, GA 30322; Department of Psychiatry, Vagelos College of Physicians and Surgeons, Columbia University. 1051 Riverside Drive, New York, NY 10032

**Keywords:** Air pollution, Particulate matter, Nitrogen dioxide, Metabolomics, Aging population, Alzheimer’s disease, Dementia

## Abstract

Long-term exposure to air pollution has been associated with changes in levels of several metabolites measured in the peripheral blood. However, most work has been conducted in ethnically homogenous populations. We studied the relationship between the plasma metabolome and long-term exposure to three air pollutants: particulate matter (PM) less than 2.5 µm in aero diameter (PM_2.5_), PM less than 10 µm in aero diameter (PM_10_) and nitrogen dioxide (NO_2_) among 107 participants of the Washington Heights and Inwood Community Aging Project (WHICAP) in New York City. Plasma metabolomic profiles were generated using untargeted liquid chromatography coupled with high-resolution mass spectrometry. We estimated the association between each metabolic feature and predicted annual mean exposure to the air pollutants using three approaches: 1. A metabolome wide association study (MWAS) framework; 2. Feature selection using elastic net regression; and 3. A multivariate approach using partial least squares discriminant analysis. Additionally, we identified the pathways enriched by metabolic features associated with exposure through pathway analysis. The samples were collected from 1995 – 2015 and included non-Hispanic white, Caribbean Hispanic, and non-Hispanic Black older adults. Through the MWAS, we found 79 features associated with exposure to PM_2.5_ (false discovery rate at 5%) but none associated with PM_10_ or NO_2_. Pathway analysis revealed that PM_2.5_ exposure was associated with altered amino acid metabolism, energy production, and oxidative stress response. Six features were found to be associated with PM_2.5_ exposure through all three approaches, annotated as: cysteinylglycine disulfide, a diglyceride, and a dicarboxylic acid. Additionally, we found that the relationship between several features and PM_2.5_ exposure was modified by diet and metabolic diseases. These signals, identified in a neighborhood-representative older population, could help understand the mechanisms through which PM_2.5_ exposure can lead to altered metabolic outcomes in an older population.

**HIGHLIGHTS:**

- Long-term exposure to PM_2.5_ is associated with altered plasma metabolic features in an aging population
- These associations are modified by a dementia diagnosis, history of diabetes, APOE-ε4 allele, and diet
- Pathways related to energy production, amino acid metabolism, and redox homeostasis are associated with exposure to PM_2.5_

**GRAPHICAL ABSTRACT:** 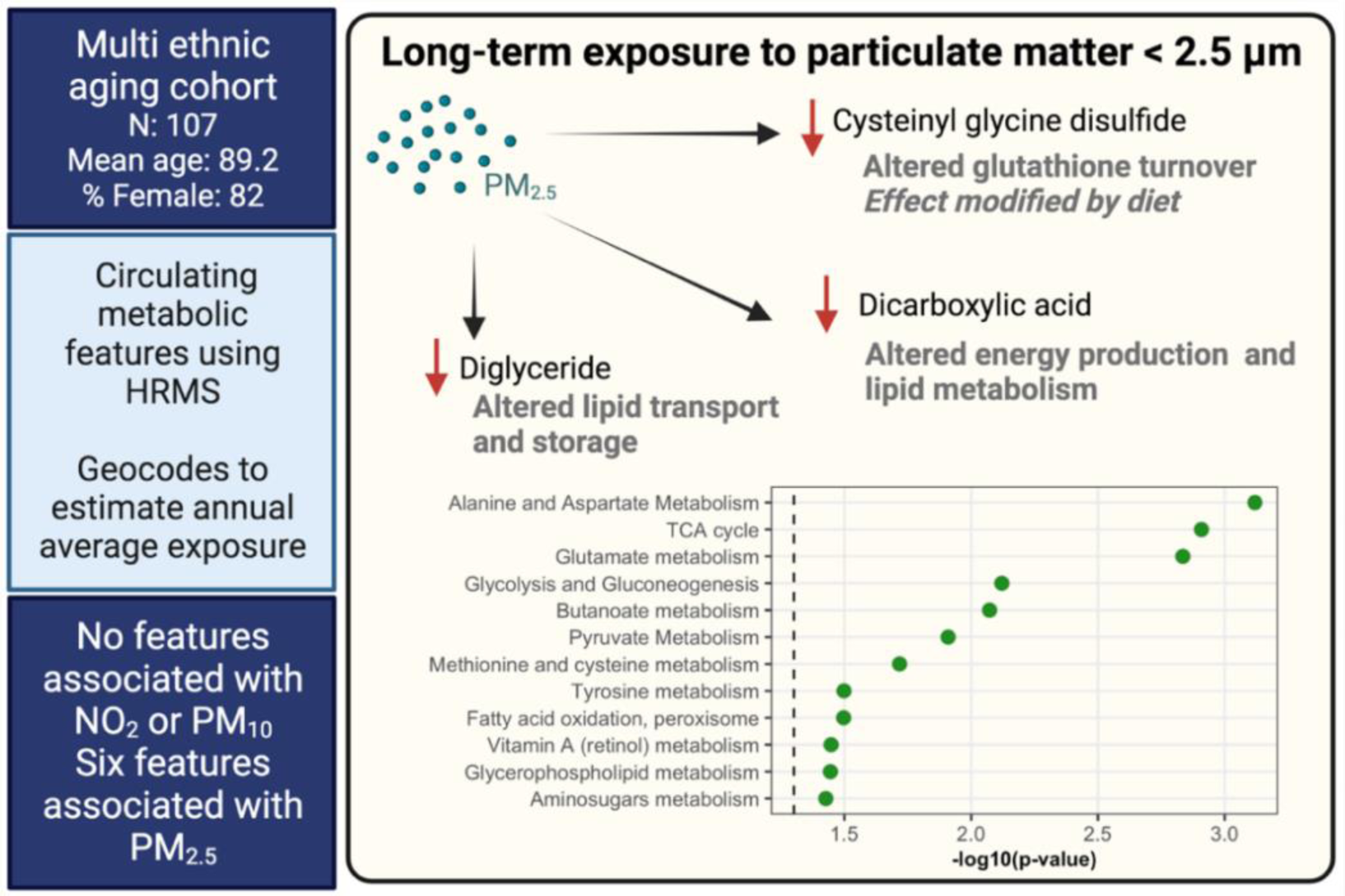

## INTRODUCTION

Air pollutants have been associated with mortality^1^ and diseases^2^ involving the pulmonary^3^, cardiovascular^4^, and nervous system^5^. These associations have been attributed to system-wide changes induced by air pollution exposure including oxidative stress^6^, inflammation^7^, and changes in circulating metabolites^8^. Long term exposure to fine particulate matter with aerodynamic diameter ≤ 2.5 µm (PM_2.5_), particulate matter with aerodynamic diameter ≤ 10 µm (PM_10_), and nitrogen dioxide (NO_2_) has been associated with decline in cognitive function^9,10^ and exposure to PM_2.5_ has been associated with clinical aggravation of age-related neurodegenerative diseases^11^.

The metabolome comprises all metabolites and other small molecules in a biological matrix that may derive from both endogenous biochemical processes, exogenous exposures that are absorbed and metabolized by the body, as well as biochemical changes that result from environmental exposures^12^. The application of high-resolution mass spectrometry-based untargeted metabolomics allows us to capture a comprehensive profile of circulating small molecules, which include both known and as yet unknown circulating small molecules^13^. Thus, by applying untargeted metabolomics, we can determine circulating small molecules associated with long-term exposure to air pollutants in an agnostic manner. A metabolic signal in the periphery as a result of exposure to air pollutants is plausible since inhaled particles have been reported to pass into circulation from the lungs^14^. Besides, particles that reach the lower airways and alveoli can induce inflammation and oxidative stress, activating local and systemic metabolic changes^15,16^. Indeed, studies have reported a circulating metabolic signature associated with exposure to traffic-related air pollutants^17^. However, few studies have investigated the relationship between long-term exposure to air pollution and circulating metabolic signals in an ethnically diverse aging population using an untargeted global metabolomic approach.

Studies have suggested that individuals with chronic diseases may be more sensitive to effects of air pollution^18^, possibly due to deficiencies in antioxidants and elevated levels of inflammation^19^. However, few studies have considered whether associations between exposure to air pollutants and circulating metabolites are modified by underlying metabolic diseases. Identifying factors that can modify this relationship would allow us to determine whether people with age-related diseases are particularly vulnerable to the effects of air pollution exposure given that people are living longer lives and bear a large burden of age-related co-morbidities^20^. Besides, older adults are particularly susceptible to the effects of chronic environmental exposures in part due to age-related deterioration in physiological function as well as the presence of co-morbidities that exert a strain on the body’s ability to respond to environmental insults^21^. One recently published study found that long-term exposure to air pollution is associated with metabolic perturbations in older adults; however, their study population only included non-Hispanic white men^22^. Disparities in exposure to air pollutants^23–25^ and mortality related to PM_2.5_ exposure^1,26^ among racial/ethnic groups has been well documented. Thus, studying the effects of air pollution exposure in an ethnically diverse aging community is necessary.

Therefore, we performed analyses to identify circulating metabolites associated with long-term exposure to PM_2.5_, PM_10_, and NO_2_ and investigated whether these relationships are modified by prevalent cardiometabolic and neurodegenerative diseases. We used data from the Washington Heights and Inwood Community Aging Project (WHICAP) in New York City, a multi-ethnic community-based cohort of older adults created to study risk factors of dementia and Alzheimer’s disease. We used three different regression methods to find reliable metabolic features associated with exposure. If any features were significantly associated (false discovery rate at 5%) with an exposure through a metabolome-wide association study framework, we additionally performed a penalized regression for feature selection, using elastic net regression, and multivariate supervised dimensionality reduction, using partial least squares differential analysis, to find metabolic features important in predicting exposure. We then report concordance of results from all three approaches. Such a multipronged approach has been used as an alternative to traditional hypothesis testing for selection of metabolic features of importance^27^. Additionally, we tested for the presence of effect modification by factors that are known to affect metabolism: sex^28^, racial/ethnic group^29^, dementia ^30^, a history of diabetes, hypertension or heart disease^31,25,32^, the presence of an APOE-ε4 allele^33^, and diet^34^. In doing so, we aimed to identify factors and morbidities that may modify the relationship between exposure and circulating metabolites.

## MATERIALS AND METHODS

### 1. Study population

We leveraged previously existing metabolomics data in 119 older adults from the WHICAP cohort^30^. WHICAP is a representative community-based cohort in the northern Manhattan region of New York city comprising of individuals aged 65 years and older enrolled through a collaborative effort with the Centers for Medicare and Medicaid and through marketing rolls. The participants were enrolled after obtaining informed consent and an interview was conducted in either English or Spanish. A detailed clinical assessment with standardized medical and neurological history was collected by a trained physician. At each visit, blood was drawn and sent to the laboratory within two hours for DNA extraction and storage of plasma and serum. At the first visit, residential information was also obtained. Results from all interviews, clinical visits, and tests were reviewed through a consensus conference comprised of clinicians with expert knowledge in the diagnosis of Alzheimer’s disease and related dementias, who made an Alzheimer’s disease diagnosis based on accepted criteria^35^. The study comprised participants equally divided among non-Hispanic white, non-Hispanic Black and Caribbean Hispanic ancestry based on self-report, classified based on the 1990 U.S. Census guidelines. Fifty-nine participants were diagnosed with clinical Alzheimer’s Disease and 60 were heathy at the time of assessment.

### 2. High resolution mass spectrometry-based metabolomics

The details on the acquisition of the untargeted metabolomics has been previously described^30,36^. The data were generated in the Clinical Biomarkers Laboratory at Emory University. Briefly, the metabolites were extracted from plasma using acetonitrile and the extracts were injected in triplicate on two chromatographic columns: a hydrophilic interaction column (HILIC) under positive ionization (HILIC+) and a C18 column under negative ionization (C18-), to obtain three technical replicates per sample per column. After separation and ionization, the ions produced were analyzed in full scan mode for molecules within 85-1250 kDa on a Thermo Orbitrap HF Q-Exactive mass spectrometer. The untargeted metabolomic data were processed through a computational pipeline that leverages open source feature detection and peak alignment software, *apLCMS*^37^ and *xMSanalyzer*^38^. The feature tables were generated containing information on the mass-to-charge (*m/z*) ratio, retention time, and the abundance/intensity of each ion for each sample. Correction for batch effects was performed using ComBat, which uses an empirical Bayesian framework to adjust for known batches in which the samples were run^39^. Each of these ions are referred to as metabolic features. For the analysis, metabolic features detected in at least 70% of the samples were retained, leaving 6375 features from the HILIC+ column and 3759 features from the C18-column for further analysis. Zero intensity values were considered below the detection limit of the instrument and were imputed with half the minimum intensity observed for each metabolic feature. The intensity of each metabolic feature was log10 transformed, quantile normalized, and auto-scaled for normalization and standardization.

### 3. Exposure assessment

The methods used for exposure assessment have been described in detail previously^9^. In brief, participants’ residential addresses obtained during the first neuropsychological examination were geocoded and long-term air pollution exposures in the calendar year prior to a clinical visit were estimated for NO_2_ (ppb), PM_10_ (µg/m^3^) and PM_2.5_, (µg/m^3^) using regionalized universal kriging models^40,41^. Measurements of the air pollutants were obtained from the US EPA Air Quality System database. For geocodes without monitoring data, the annual averages were used in the kriging models to predict concentrations. To improve predictions, information on land use, distance to roadways, and other geographic variables were included using partial least square methods. The exposure data were analyzed as continuous variables in linear and penalized regressions and dichotomized for the partial least square discriminant analysis. In this study we considered 107 participants for whom both metabolomics data and air pollution exposure estimates were available.

### 4. Statistical analysis

We used a metabolome-wide association study (MWAS) framework with correction for multiple comparisons by controlling the false discovery rate (FDR) at 5%. In case significant associations between metabolic features and an air pollutant were observed we then performed two additional tests, elastic net regression (e-net) and partial least squares discriminant analysis (PLS-DA). The overlap of metabolic features in the results from all three analyses was determined using the *VennDetail* package (version 1.14.0) which extracts details of multi-set interactions that can be used for visualization. All analyses were conducted in R (version 3.6.3).

#### 4.1. Metabolome-wide association with exposure to air pollutants and pathway analysis

We estimated the relationship between estimated long-term exposure to PM_2.5_, PM_10_, and NO_2_ and each metabolic feature using multiple single-pollutant models, adjusted for potential confounders and predictors of the outcome including age (years), sex (women/men), racial/ethnic group (non-Hispanic white, non-Hispanic Black, Caribbean Hispanic), clinical diagnosis of Alzheimer’s disease (case/control (no dementia)), year of blood draw, and years of education. The analyses were conducted separately for data from each column. We corrected for multiple comparisons using an FDR of 5% and q-values were estimated using the Benjamini-Hochberg (BH) method.

#### 4.2. Penalized regression for selection of metabolic features that predict exposure to PM_2.5_

The MWAS only detected significant associations with PM_2.5_; we thus, performed elastic net regression to determine which features from both columns were predictors of PM_2.5_ exposure. The covariates included were the same as the MWAS framework. We used five-fold cross-validation, repeated five times, using the *caret* package (version 6.0-90) with a custom grid search where alpha ranged from 0.1 to 1 with 0.1 increments, and lambda ranged from 0.0001 to 1 with 0.053 increments, to determine the optimal penalization and mixing parameters^42^. Based on the root mean square error, the parameters chosen were a lambda of 0.158 and alpha of 1 (i.e., lasso was the optimal fit).

#### 4.3. Partial least squares discriminant analysis

Finally, we conducted a partial least squares discriminant analysis to determine which metabolic features were important in discriminating between low and high exposure to PM_2.5_ using a multivariate approach. We created a binary categorization of the PM_2.5_ exposure by first calculating the residuals from regressing PM_2.5_ exposure on year of blood draw, since there was a strong negative correlation between PM_2.5_ exposure and year of blood draw. Second, we categorized exposure as high or low at the median of the residuals. Then, we regressed the intensity of each metabolic feature from both columns on the same covariates as in §4.1 and stored the residuals. Finally, we used the de-trended metabolic feature intensities in a PLS-DA model using the *mixOmics* package (version 6.11.1), which contains methods for multivariate methods for omics data^43^. We used a variable importance VIP score cut-off of 3.0 for component 1 and 2.5 for component 2, to deem a metabolic feature important in discriminating between high and low exposure. These cut-offs were chosen to find the most important features based on the distribution of VIP scores.

#### 4.4. Pathway analysis

To determine the biological relevance of the metabolic features associated with PM_2.5_, we conducted pathway analysis using the “functional analysis” module in *MetaboAnalyst* (version 5.0)^44^, a web-based interface for comprehensive metabolomic data analysis. We used the MWAS results from both columns and applied a nominal p-value cut-off of 0.01 to determine metabolic pathway enrichment using the mummichog algorithm and the human MFN reference database^45^. We present results for pathways with a Fisher’s exact test p-value < 0.1.

#### 4.5. Metabolite annotation

Metabolite annotations were made using an internal library and by matching to the Human Metabolome Database (HMDB)^46^, the Kyoto Encyclopedia of Genes and Genomes (KEGG) database, and LIPIDMAPS using the R package *xMSannotator* (version 1.3.2) ^47^. This uses a multistage clustering algorithm method to determine metabolic pathway associations, intensity profiles, retention time, mass defect, and isotope/adduct patterns to assign putative annotations to metabolic features. In cases where a feature had multiple matches, we used the following rules to assign an annotation: first, we screened features based on the confidence score assigned by xMSannotator, and the annotation with the highest score was used. Second, if all annotations had the same score, we chose the annotation with the lowest difference in expected and observed mass (delta ppm). Finally, if all features had the same score and delta ppm, we indicated the identity as “multiple matches” since we couldn’t decipher a unique putative annotation. The confidence in annotation was based on criteria defined by Schymanski et al^48^, where level 1 corresponds to a confirmed structure identified through MS/MS and/or comparison to an authentic standard; level 2 to a probable structure identified through spectral matches to a database; level 3 to a putative identification with a speculative structure; level 4 to an unequivocal molecular formula but with insufficient evidence to propose a structure; and level 5 to an exact mass but not enough information to assign a formula.

#### 4.6. Effect modification and sensitivity analysis

We tested for the presence of effect modification only in the metabolic features that were significantly associated with long-term exposure to PM_2.5_ after correction for multiple comparisons through the MWAS approach. This 2-step approach that utilizes a screening step has been shown to improve power to detect gene-by-environment interactions^49^. We compared models with and without an interaction term between PM_2.5_ and the effect modifier using likelihood ratio tests and controlled the FDR at 20% to look for noteworthy interactions using the Benjamini-Hochberg method.

##### 4.6.1.

We tested whether racial/ethnic group, sex, history of heart disease, hypertension or diabetes, an Alzheimer’s disease diagnosis, or an APOE-ε4 allele were effect modifiers of the observed associations. We ran separate linear models for each modifier, adjusted for same covariates as in §4.1. and included an interaction term for each modifier and PM_2.5_. This model was compared to a lower-order model with the modifier included only as a covariate, without an interaction term.

##### 4.6.2.

To test for effect modification by diet we used data from a 61-item version of the Willet semiquantitative food frequency questionnaire (SFFQ). This questionnaire has been validated for use in older adults^50^ and in the WHICAP cohort^51^. The questionnaire was administered in English or Spanish to participants of the WHICAP cohort to determine average food consumption in the year prior to baseline assessment. The 61 food items were categorized into 30 groups and intake of each group was calculated by summing intakes of member food items. The daily intake of nutrients was determined by multiplying the frequency of consumption of each portion of every food by the nutrient content of each portion^52,53^. In our study, we had the SFFQ data available for 77 of the observations (72%). We used data on estimated intake of four macronutrients: total carbohydrate, total protein, animal fat, and vegetable fat. The estimated intake of the macronutrients was adjusted for total caloric intake using the residual method. We used principal component analysis to create linear combinations of the four macronutrients to discern patterns in macronutrient intake using the *prcomp()* function in R. Each of the principal components (PC) was used to test for effect modification by including an interaction term between each PC and PM_2.5_ in a linear model like that in §4.6.1. and additionally, adjusting for total caloric intake.

##### 4.6.3.

Sensitivity analysis was conducted to determine whether smoking history confounded the relationship observed between long-term PM_2.5_ exposure and the metabolic features by additionally adjusting the model in §4.1. for smoking history. The results were compared by plotting the coefficients for PM_2.5_ from the two models to determine whether adjusting for smoking history produced coefficients different from the main analysis.

## RESULTS

The mean age of the participants was 89.2 years (standard deviation (SD) = 8.44), women made up 82% of the population, 34.6% were non-Hispanic Black, 33.6% were non-Hispanic white, and 31.8% were Caribbean Hispanic, and the mean number of years of education was 9.42 (SD = 4.66). Roughly half of the participants were diagnosed with Alzheimer’s disease (53.3%), 27.1% of the participants carried at least one APOE-e4 allele, 20.6% of the participants had a history of diabetes, 37.4% had a history of heart disease, and 71% had a history of hypertension. The metabolomic data consisted of 6375 metabolic features from the HILIC+ column and 3759 features from the C18-column after data filtering and cleaning. The average long-term exposures in these participants was 31.7 (SD = 7.03) ppb for NO_2_, 21.0 (SD = 7.73) µg/m^3^ for PM_10_ and 12.9 (SD = 2.41) µg/m^3^ for PM_2.5_ (Table 1).

**Table 1.** Characteristics of the study population in the Washington Heights and Inwood Community Aging Project. After regressing out the effect of year of blood draw, the PM_2.5_ exposure levels were dichotomized at the median, to create low and high exposure groups. This was also used to determine features associated with exposure through a partial least square discriminant analysis.

|  | Low PM<sub>2.5</sub><br>(N=54) | High PM<sub>2.5</sub><br>(N=53) | Overall<br>(N=107) |
| --- | --- | --- | --- |
| **Age (years)** |  |  |  |
| Mean (SD) | 90.7 (7.31) | 87.6 (9.25) | 89.2 (8.44) |
| **Sex** |  |  |  |
| Women | 44 (81.5%) | 44 (83.0%) | 88 (82.2%) |
| Men | 10 (18.5%) | 9 (17.0%) | 19 (17.8%) |
| **Racial/Ethnic group** |  |  |  |
| Non-Hispanic Black | 22 (40.7%) | 15 (28.3%) | 37 (34.6%) |
| Non-Hispanic white | 16 (29.6%) | 20 (37.7%) | 36 (33.6%) |
| Caribbean Hispanic | 16 (29.6%) | 18 (34.0%) | 34 (31.8%) |
| **Education (years)** |  |  |  |
| Mean (SD) | 9.74 (4.81) | 9.09 (4.53) | 9.42 (4.66) |
| **Alzheimer's disease** |  |  |  |
| Case | 28 (51.9%) | 29 (54.7%) | 57 (53.3%) |
| Control | 26 (48.1%) | 24 (45.3%) | 50 (46.7%) |
| **APOE-ε4 carrier** |  |  |  |
| No ε4 allele | 39 (72.2%) | 39 (73.6%) | 78 (72.9%) |
| At least one ε4 allele | 15 (27.8%) | 14 (26.4%) | 29 (27.1%) |
| **History of diabetes** |  |  |  |
| No | 45 (83.3%) | 40 (75.5%) | 85 (79.4%) |
| Yes | 9 (16.7%) | 13 (24.5%) | 22 (20.6%) |
| **History of heart disease** |  |  |  |
| No | 36 (66.7%) | 31 (58.5%) | 67 (62.6%) |
| Yes | 18 (33.3%) | 22 (41.5%) | 40 (37.4%) |
| **History of hypertension** |  |  |  |
| No | 21 (38.9%) | 10 (18.9%) | 31 (29.0%) |
| Yes | 33 (61.1%) | 43 (81.1%) | 76 (71.0%) |
| **PM<sub>2.5</sub> (μg/m<sup>3</sup>)** |  |  |  |
| Mean (SD) | 11.8 (2.65) | 14.0 (1.48) | 12.9 (2.41) |
| **PM<sub>10</sub> (μg/m<sup>3</sup>)** |  |  |  |
| Mean (SD) | 20.7 (8.82) | 21.2 (6.51) | 21.0 (7.73) |
| **NO<sub>2</sub> (ppb)** |  |  |  |
| Mean (SD) | 29.8 (7.96) | 33.7 (5.29) | 31.7 (7.03) |

Using an MWAS framework, we found 79 metabolic features significantly associated with PM_2.5_ (61 from the HILIC+ column and 18 from the C18-column, Figure 1, Supplemental table 2) after correcting for multiple comparisons. No significant associations were found between the metabolic features and PM_10_ or NO_2_ (Figure 1). Through the penalized elastic net regression, we found 33 metabolic features that had non-zero coefficients (19 from HILIC+ column and 16 from C18-, Supplemental table 3). The results from the PLS-DA showed separation along component 1 and 2 of people exposed to PM_2.5_ below or above the median (Figure 2A). Thirty-six metabolic features had a variable importance score > 3 on component 1 or a VIP score > 2.5 on component 2 (32 from HILIC+ column and 4 from C18-column, Supplemental table 4).

**Figure 1.**
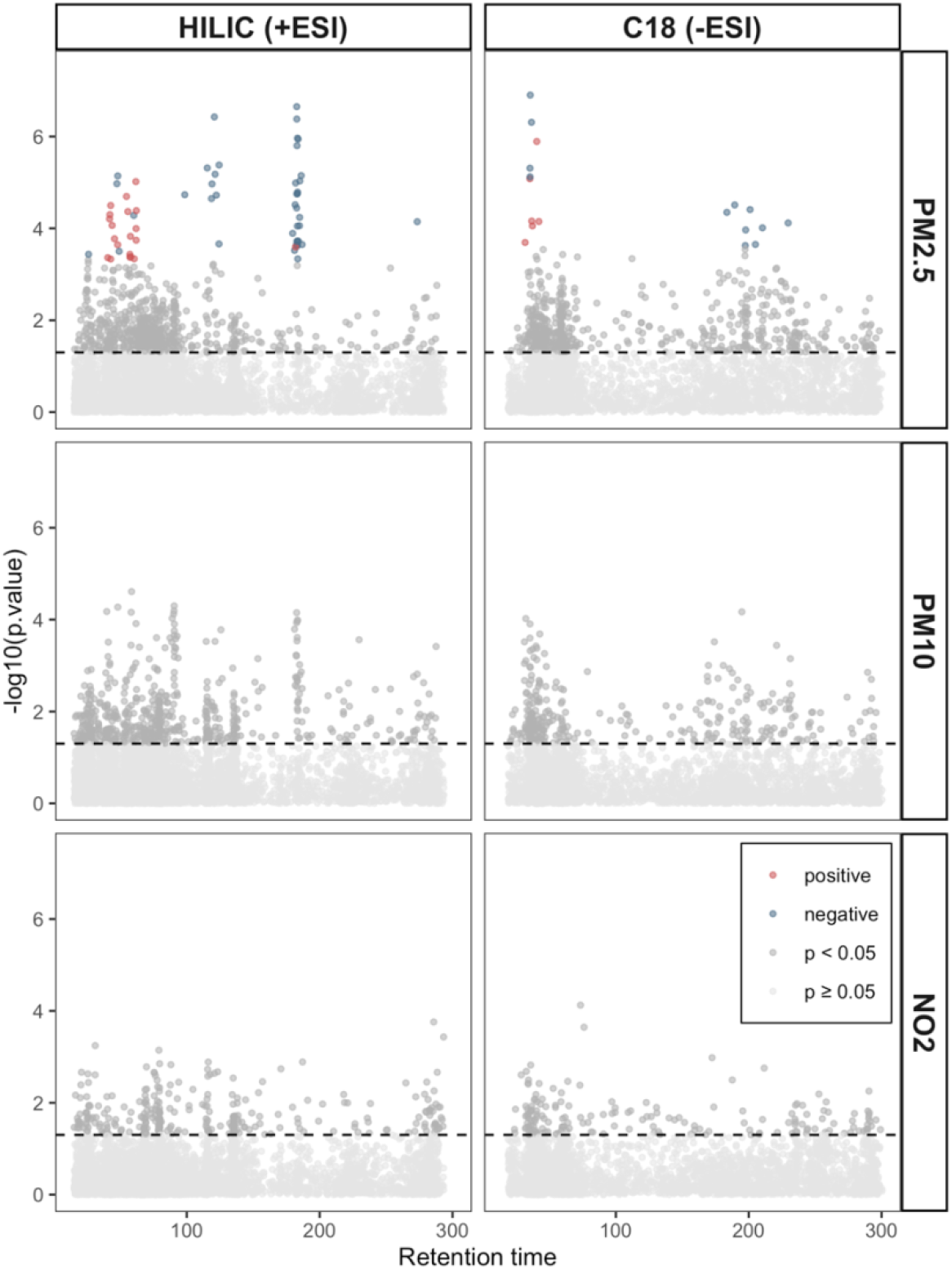
Manhattan plots show the metabolic features associated with long-term exposure to air pollutants determined through a metabolome wide association study framework. In the grid, the two columns represent data from the HILIC (on the left) and C18 chromatographic column under positive and negative ionization respectively. The rows show data for the respective air pollutants. All models were adjusted for age (years), sex (men/women), racial/ethnic group (non-Hispanic white, non-Hispanic black, Hispanic Caribbean), Alzheimer’s disease status (case/control), year of blood draw and years of education. The horizontal line represents nominal p-value < 0.05. The colored dots represent features significantly associated with the pollutant after correction for multiple comparisons (FDR < 0.05) with blue dots representing a negative association between that metabolic feature and air pollutant and the red dots a positive association.

**Figure 2.**
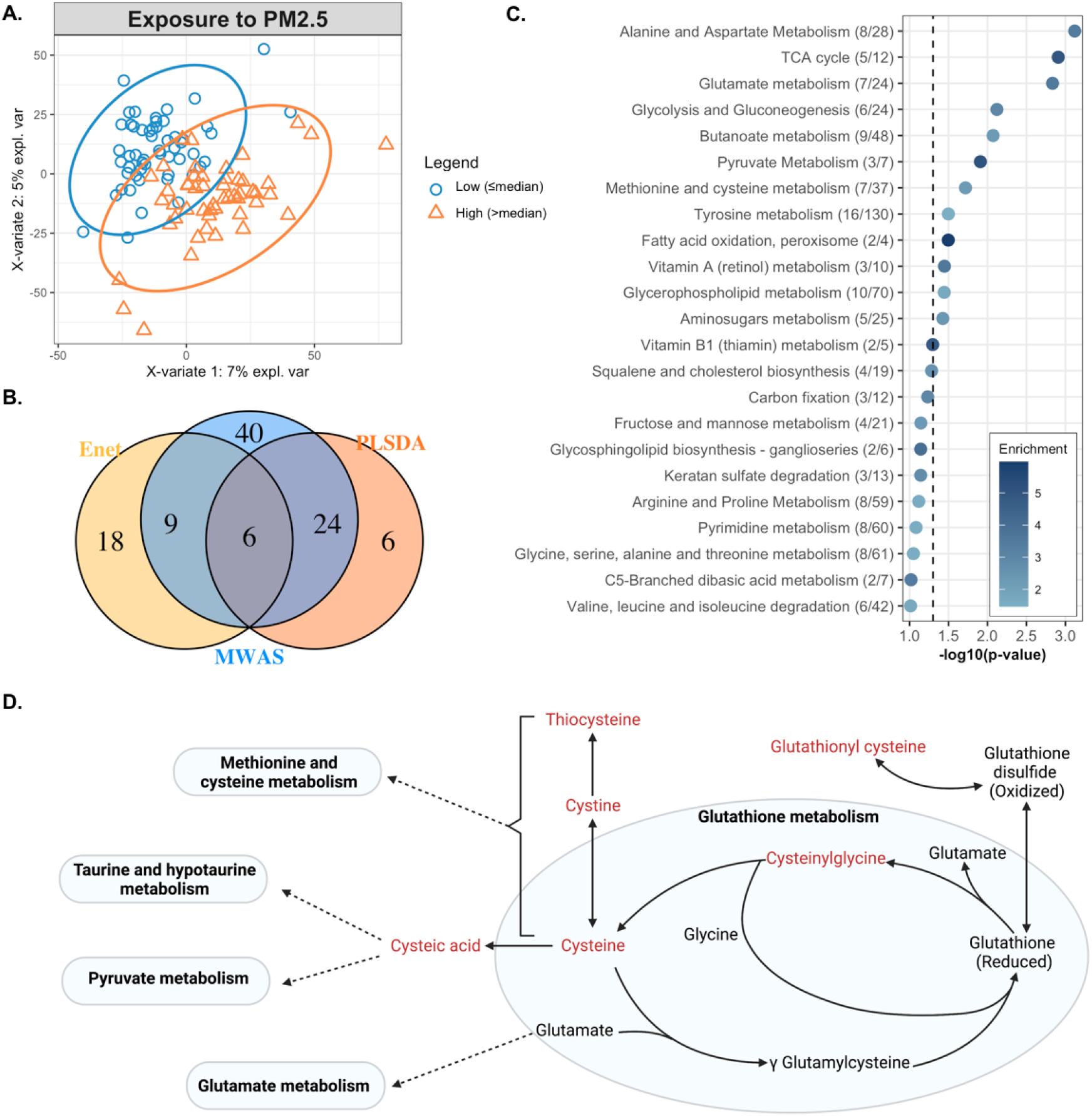
In A, the biplot shows clustering of observations with high (orange triangles) and low (blue circles) long-term exposure to PM_2.5_ after adjusting for age, sex, ethnic group, Alzheimer’s disease status, year of blood draw and years of education. In B, the overlap in metabolic features associated with long-term PM_2.5_, estimated through three different approaches, a metabolome association study (MWAS), elastic net regression (enet) and partial least squares discriminant analysis (PLSDA). In C, the metabolic pathways enriched by the metabolic features associated with long-term PM_2.5_ exposure, determined through the MWAS approach. Enrichment represents that ratio between observed number of significant hits/expected number of hits. In D, a representation of glutathione metabolism and the biochemical relationship between members of glutathione metabolism and other pathways. Metabolites in red text were negatively associated with PM_2.5_ through either MWAS, enet, or PLS-DA.

When comparing the results from the three approaches, we found 6 features associated with PM_2.5_ through all approaches, i.e., the MWAS, elastic net and PLS-DA (Figure 2B). These features were putatively annotated as cysteinyl glycine disulphide, a diglyceride, alphachloralose, a dicarboxylic acid, and one feature had multiple matches in HMDB (Table 2). The metabolic features associated with PM_2.5_ enriched several metabolic pathways including: alanine and aspartate metabolism, the TCA cycle, glutamate metabolism, glycolysis and gluconeogenesis, butanoate metabolism, pyruvate metabolism, methionine and cysteine metabolism, tyrosine metabolism, fatty acid oxidation, vitamin A metabolism, glycerophospholipid metabolism, and aminosugars metabolism. These represented pathways related to energy production, redox homeostasis, and amino acid metabolism (Figure 2C).

**Table 2.** The putative annotations of the six features associated with long-term PM_2.5_ through all three approaches (Figure 2 B). m/z: mass-to-charge ratio, Time: Retention time, Delta ppm: mass difference in parts per million, ID score: confidence in annotation based on Schymanski scale (1 being the highest and 5 the lowest), ESI: electrospray ionization

| m/z | Time (s) | Annotation | Formula | Delta ppm | Adduct | ID score* | ESI | $\beta$ | p-value | Pathway/source |
| --- | --- | --- | --- | --- | --- | --- | --- | --- | --- | --- |
| 300.0478 | 120.7 | Cysteinylglycine disulfide, multiple other matches | C8H15N3O5S2 | 0 | M+H [+2] | 3 | + | -0.48 | 3.7E-07 | Glutathione metabolism; Redox homeostasis |
| 149.53 | 124.3 | Cysteinylglycine disulfide | C8H15N3O5S2 | 0.47 | M+2H | 3 | + | -0.45 | 4.2E-06 | Glutathione metabolism; Redox homeostasis |
| 355.0733 | 115.4 | Multiple matches | - | - | - | 5 | + | -0.44 | 4.8E-06 | - |
| 154.9871 | 183.3 | Alphachloralose | C8H11Cl3O6 | 8 | M+2H | 5 | + | -0.45 | 1.1E-06 | Rodenticide |
| 775.6017 | 48 | A diglyceride | C45H72O5 | 4.32 | M+2ACN+H | 3 | + | -0.43 | 7.2E-06 | Lipid metabolism |
| 129.0194 | 36.5 | A dicarboxylic acid | C5H6O4 | 0.54 | M-H | 3 | - | -0.49 | 4.9E-07 | Lipid metabolism |

None of the relationships identified as significant through MWAS were modified by sex, racial/ethnic group, history of heart disease, or history of hypertension. The relationship between a lysophosphatidylethanolamine and PM_2.5_ was modified by both Alzheimer’s disease diagnosis and APOE-ε4 allele status, such that people with Alzheimer’s disease or carriers of at least one ε4 allele had a negative association between exposure and levels of lysophosphatidylethanolamine while those without Alzheimer’s disease or an ε4 allele had a positive association (Table 3, Supplemental figure 1). The relationship between two other features annotated as lysophosphatidylethanolamine was also modified by APOE-ε4 allele status in a similar way, i.e., those with at least one ε4 allele had a negative association between exposure and lysophosphatidylethanolamine (Table 3, Supplemental figure 2). The relationship between three features and PM_2.5_ was modified by a history of diabetes, such that people with a history of diabetes had a greater positive association between the features and exposure to PM_2.5_ (Table 3, Supplemental figure 3), however; the features did not match any database entries.

**Table 3.**
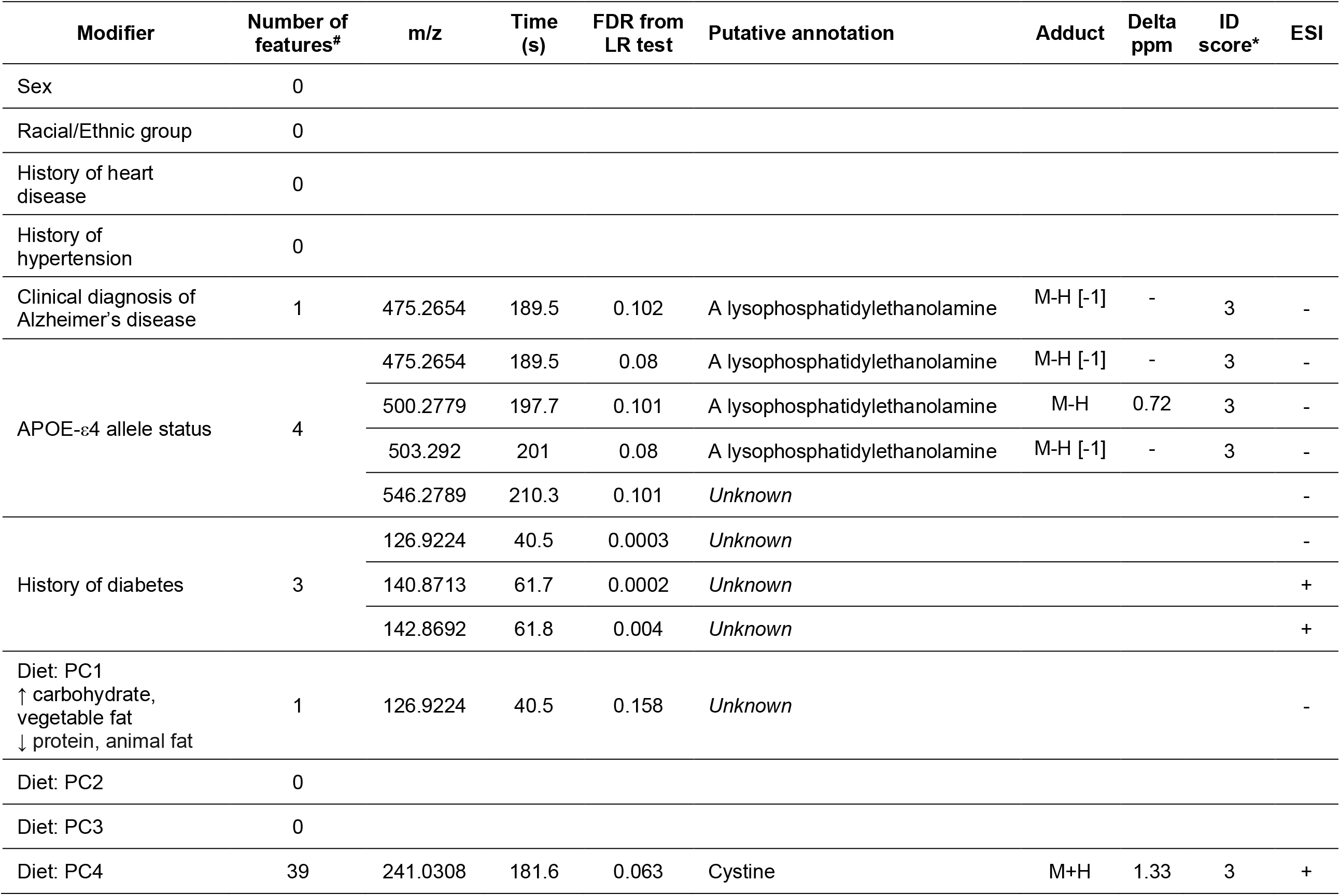
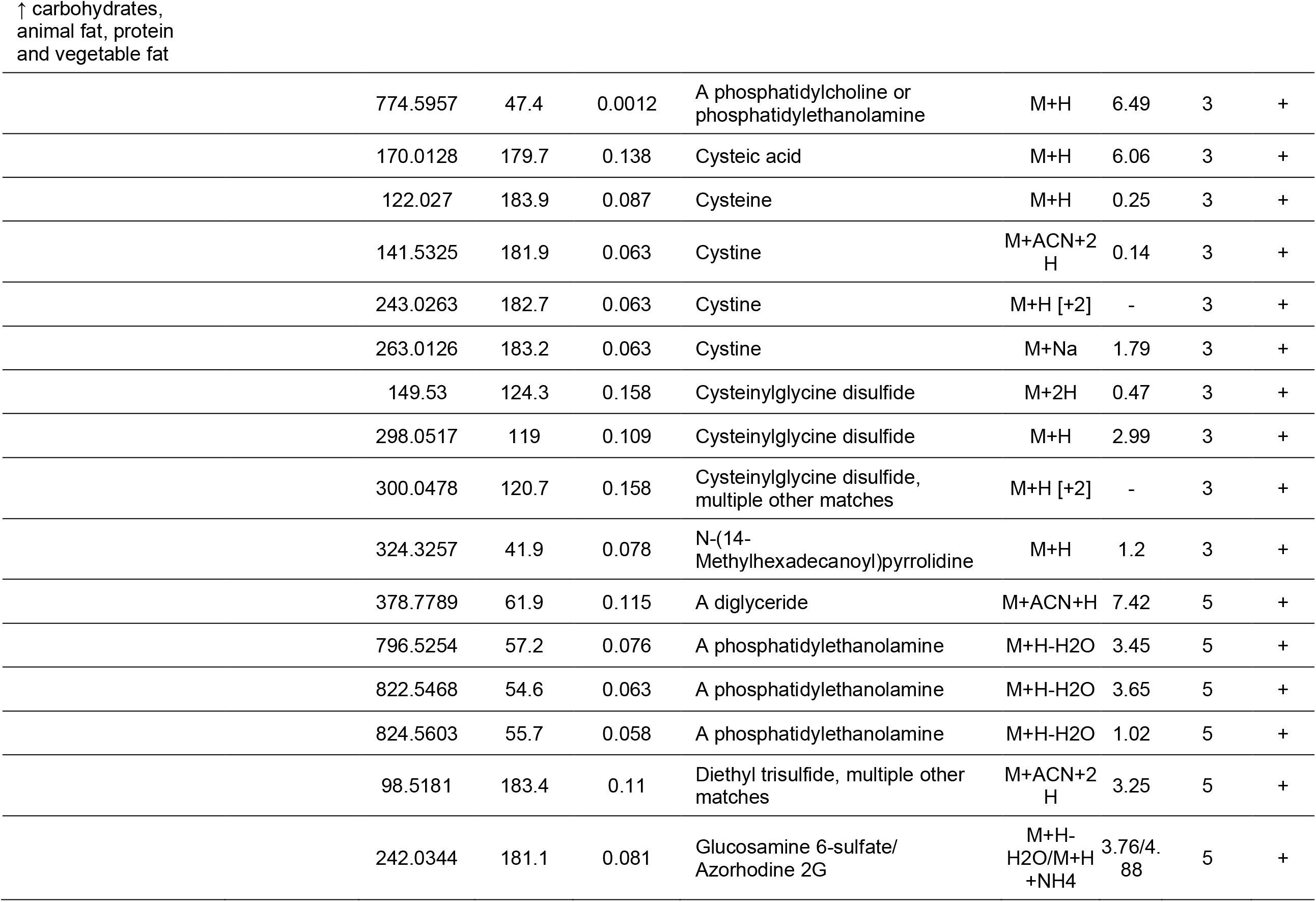
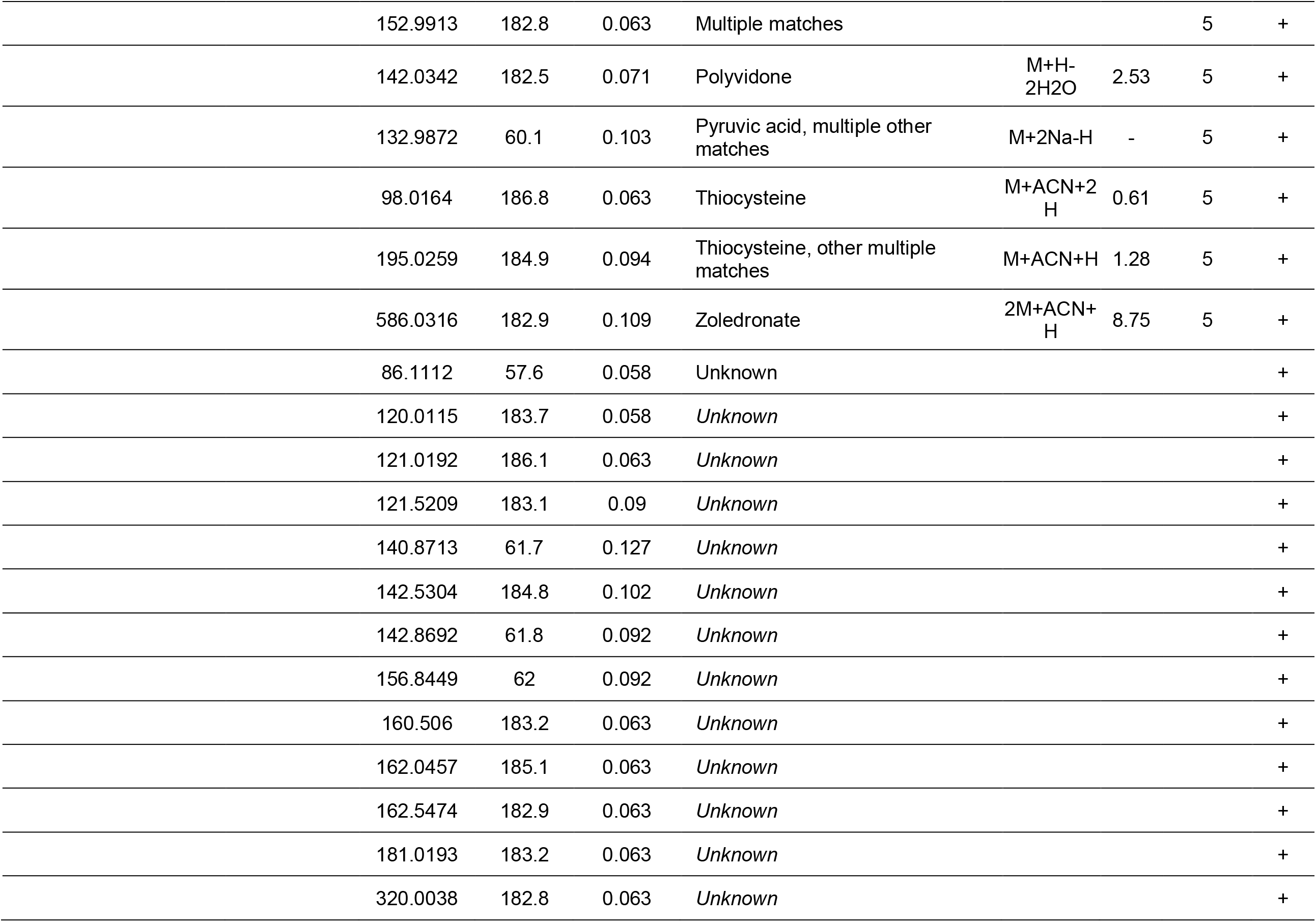
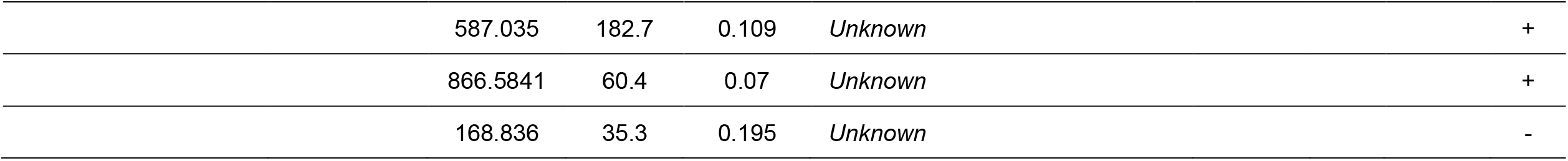
Effect modification. The relationship between several metabolic features and PM_2.5_ was modified by Alzheimer’s disease diagnosis, APOE-ε4 allele, history of diabetes, and diet. ^#^ A noteworthy interaction was defined at an FDR < 0.2. LR: Likelihood ratio test, m/z: mass-to-charge ratio, Time: retention time, Delta ppm: mass difference in parts per million, ID score: confidence in annotation based on Schymanski scale (1 being the highest and 5 the lowest), ESI: electrospray ionization.

The principal component analysis of macronutrients from the dietary data generated four PCs (Supplemental figure 4). PC1 (explained 57.1% of the total variance in the data) had positive loadings from carbohydrates and vegetable fat and negative loadings from total protein and animal fat and modified the relationship between one metabolic feature and exposure to PM_2.5_ such that a unit increase in the score on PC1 reduced the slope of the relationship between the metabolic feature and PM_2.5_. PC2 (28% of variance explained) and PC3 (13.9% of variance explained) did not modify any relationships discovered but the relationship between 39 of the metabolic features, about half of the features identified in MWAS, and PM_2.5_ was modified by PC4 (explained 0.01% of the variance in the data), which had positive loadings from all four macronutrients (Table 3, Supplemental figure 4).

In sensitivity analysis, the associations between exposure and the metabolic features were not affected by the inclusion of smoking history as a covariate in the model (Supplemental figure 5).

## DISCUSSION

We found several plasma metabolic features associated with predicted outdoor residential annual concentrations of PM_2.5_ in a racially and ethnically diverse urban population of older adults. Pathway analysis revealed perturbations in the metabolism of several amino acids, the citrate cycle, glycolysis and gluconeogenesis, butanoate, pyruvate, fatty acid, vitamins and co-factors, glycerophospholipid, and aminosugars. These metabolic pathways have been previously associated with PM_2.5_ and include indicators of oxidative stress. These signals could help us understand the mechanisms through which PM_2.5_ exposure can lead to altered health outcomes.

This is the first study of metabolic features associated with long-term exposure to PM_2.5_ in a racially and ethnically diverse aging population of both men and women. Additionally, we used three different statistical approaches to determine features associated with exposure to PM_2.5_. By looking for concordance among results from diverse regression methods, we found a reliable set of features that are associated with, and can predict exposure to, PM_2.5_. While others have used such multipronged approaches for metabolomic data analysis,^27^ this is the first study to consider such an approach when studying the effect of long-term exposure to PM_2.5_ on the circulating metabolome. We identified six features associated with PM_2.5_ through this approach, two of which were annotated as cysteinylglycine disulfide (an M+2H adduct and an isotope of the M+H adduct), one as dicarboxylic acid, and another as diglyceride with level 3 confidence, and two were annotated with level 5 confidence, one as alphachloralose and another with multiple database matches.

Both features annotated as cysteinylglycine disulfide (cys-gly) were negatively associated with exposure to PM_2.5_. A previous study in the Normative Aging Study also reported a significant negative relationship between cys-gly and long-term exposure to PM_2.5_^22^. Cys-gly is formed by the degradation of reduced glutathione metabolism catalyzed by glutathione specific γ-glutamylcyclotransferase^54,55^. This cleavage of reduced glutathione is important for the turnover of glutathione (Figure 2D), which is required for maintaining redox homeostasis^55^, and in cellular stress response^56^. A reduction in glutathione levels has also been observed in the liver of mice exposed to PM_2.5_ ^57^.

Dicarboxylic acids are produced by breakdown of amino acids and fatty acids and can feed into the TCA cycle for energy production^58,59^. The observed association may suggest changes in the energy production from amino acids and fatty acids because of long-term exposure to PM_2.5_. In fact, pathway analysis did reveal alterations in pathways related to energy production, namely the TCA cycle and glycolysis and gluconeogenesis. *In vivo* exposure to PM_2.5_ in mice also showed altered TCA cycle, glucose, and lipid metabolism in the liver^57^.

Diglycerides (DG) consist of a glyceride with two fatty acid chains. DGs serve as substrates to the diacylglycerol acyltransferase (*Dgat*) enzyme, which converts DGs to triglycerides, metabolites that are key regulators of lipid transport and deposition in adipose tissue^60,61^. Mice exposed to PM_2.5_ had higher expression of *Dgat* in white adipose tissue^62^, suggesting an increase in the synthesis of triglycerides from DGs. This response to exposure to PM_2.5_ would also be in line with the negative association between the circulating DG and PM_2.5_ observed in our population.

Finally, two features had low confidence in their annotation: one was putatively annotated as alphachloralose while the other had multiple database matches. Given the low confidence in annotation, we do not discuss their potential role in association with PM_2.5_ but plan to confirm the identity of these features in the future.

Several of our findings confirm results from previous analyses. Long-term exposure to PM_2.5_ over 2000-2016 in an aging population of non-Hispanic white men living in the Boston area, with mean age of 75 years, was also associated with alterations in glycerophospholipid, alanine, and glutathione metabolism^22^. Similar pathways were also reported to be altered by PM_2.5_ exposure in a subset of the UKTwins study, including pyruvate metabolism, glycolysis, and gluconeogenesis^63^. In a study of younger individuals, long-term exposure to PM_2.5_ was associated with circulating phospholipids, similar to the enrichment in glycerophospholipid metabolism in our study^64^. Nearly all the metabolomic pathways affected by long-term PM_2.5_ in our study were also reported in a study of exposure to ultrafine particulate matter^65^. Additionally, animal and *in vitro* studies have also reported similar changes in metabolic pathways. Changes in the TCA cycle, amino acid biosynthesis, and glutathione metabolism were reported in lung epithelial cells exposed to fine particulate matter^66^. In mice chronically exposed to ambient PM_2.5_, serum metabolomic analysis revealed changes in glycerophospholipid, sphingolipids, glycerolipids and lysophospholipids, and in pathways related to protein digestion and absorption, glycine, serine, threonine metabolism, alanine metabolism, carbon metabolism, saccharides, fatty acids, sterols, and stress hormones^67^.

Several studies of short-term PM_2.5_ exposure have reported changes in arginine, histidine, linoleate, and leukotriene metabolism^17,65,68,69^. We did not find significant changes in levels of these metabolites associated with long-term exposure to PM_2.5_ in our study, suggesting that there might be exposure-window specific changes in circulating metabolites.

Through our analysis, we also wanted to discover whether the associations identified between circulating metabolites and exposure to long-term PM_2.5_ were modified by sex, racial/ethnic group, underlying metabolic diseases (history of diabetes, dementia diagnosis, history of heart disease, history of hypertension), or diet. The relationship between two of the six features associated with PM_2.5_, identified through all three statistical approaches, both annotated as cysteinylglycine disulfide, was modified by diet. None of the other possible modifiers affected the associations between the six features and exposure.

APOE-ε4 is a known risk factor for Alzheimer’s disease, the gene may underlie the observed modification by the dementia diagnosis. The APOE-ε4 gene plays a role in transporting cholesterol and other fats in the bloodstream, especially in the brain^70^. Lysophosphatidylethanolamine (LysoPE) is a derivative of phosphatidylethanolamine, a phospholipid typically found in the cell membrane, especially of the central nervous system^71^. Our findings suggest that people with at least one APOE-ε4 allele may have a different response to long-term exposure to PM_2.5_ possibly due to altered lipid transport. Studies in APOE null mice have reported increased lipid deposition when exposed to concentrated PM2_.5_^72^ and altered atherosclerotic plaque formation upon diesel exhaust exposure^73^ compared to mice that were unexposed.

Having a history of diabetes modified the relationship between PM_2.5_ and three features. These features did not match any entries in HMDB, KEGG, or LIPIDMAPS. However, one of these features was also modified by dietary PC1, suggesting the feature may derive from food or some other exogenous source.

Several studies have reported that the relationship between exposure to air pollutants and health outcomes is modified by diet^74^. For example, vitamin C supplementation has been shown to protect from adverse effects of ozone^63,75,76^. Similarly, a Mediterranean diet has been shown to modify the risk of long-term exposure to PM_2.5_ and cardiovascular disease mortality^77^. Consumption of animal-based food also modified the relationship between exposure to air pollution and gestational diabetes^78^, while a plant-based diet has been shown to protect against adverse effects of PM_2.5_ on cognitive function^79^.

We used macronutrient data to find whether broad changes in dietary intake influenced any of the relationships identified through the MWAS approach. To capture patterns in the macronutrient data, we used principal component analysis. We found that one feature was modified by PC1, which captured the most variance in the data, had positive loadings from carbohydrates and vegetable fat, and had negative loadings from protein and animal fat, suggesting a diet with low protein and animal-based products. However, this feature did not match entries in HMDB, KEGG, or LIPIDMAPS. Nearly half of the features that were significantly associated with PM_2.5_ had a significant interaction with PC4, which captured <1% of the variance in the dietary data and had positive loadings from all four macronutrient categories. This finding suggests that macronutrient intake modifies the relationship between circulating metabolites and long-term exposure to PM_2.5_, albeit only a small variation in the macronutrient intake data was responsible for this. It could also suggest that differences in the intake of nutrients other than the four macronutrients may modify the relationship between these metabolic features and PM_2.5_. Analysis using additional dietary data will be needed to confirm these findings.

We would like to acknowledge some limitations in the analyses presented here. First, due to our small sample size we may have been underpowered to discover associations with small effect sizes. This could explain why we found no associations between circulating metabolites and long-term exposure to PM_10_ and NO_2,_ which others have reported^22,80,81^. Second, we could only provide level 3 confidence in metabolite identification. The accurate identity of these metabolites will be determined as we confirm more metabolites on our platform using chemical standards. Third, due to the small sample size we were likely underpowered to detect significant interactions, especially with the dietary data, for which we only have 77 observations (∼72% of the total study sample). Fourth, the estimates of air pollution exposure were predicted for each participant at baseline and maybe prone to exposure measurement error. Additionally, the estimates did not account for time spent outside of the home, thus we are unable to account for exposures experienced in geographical locations outside the home, like exposures in occupational settings. However, since most of the participants were retired at the time of the study, occupational exposure is unlikely to confound the relationships observed here. Fifth, since the samples were selected for equal representation of racial/ethnic groups and Alzheimer’s disease status, we may be prone to selection bias; however, the mean levels of air pollutant exposure were similar between Alzheimer’s disease cases and controls (Supplemental table 1). Sixth, our study included people with and without Alzheimer’s disease but is not a case-control study which may reduce the generalizability of our study to the general population. Finally, since we were interested in discovering whether presence of metabolic diseases, APOE-ε4 allele and diet can modify the relationships observed, we used an FDR threshold of 20%, which may be considered lenient and not robust to false positive associations. However, we chose this threshold given the exploratory nature of our question and small sample size, and we believe the findings of effect modification warrant further exploration in larger studies.

Despite these limitations, our study has several strengths. Our sample was representative of the neighborhoods in northern Manhattan. We applied high-resolution mass spectrometry-based metabolomics to capture the circulating metabolome in an agnostic manner. Additionally, we had information on history of chronic disease with metabolic consequences and dietary information that has been well validated in the cohort. This study is also a critical step as we expand our analysis to the larger WHICAP cohort.

## CONCLUSION

Using an untargeted metabolomics analysis, we applied a multipronged approach to find circulating biochemical signals associated with estimated long-term exposure to PM_2.5_. We found that circulating cysteinylglycine disulfide, a diglyceride, and a dicarboxylic acid are associated with long-term exposure to PM_2.5_, suggesting changes in glutathione turnover, lipid transport and storage, and energy production. We also found changes in several metabolic pathways, that have previously been associated with long-term exposure to PM_2.5_. The relationship between long-term PM_2.5_ and cysteinylglycine disulfide was modified by diet suggesting that dietary changes may influence this relationship. In the future, we plan to identify the features putatively annotated in this analysis. Finally, we plan to investigate the relationship between long-term exposure to air pollutants and circulating metabolites in the larger WHICAP study. This will provide more power to also consider the relationship between long-term exposure to NO_2_ and PM_10_ and explore the role of diet in the relationship between long-term exposure to air pollutants and circulating metabolites.

## Data Availability

All data produced in the present study are available upon reasonable request to the WHICAP investigators.

## ACKNOWLEDGEMENTS

Data collection and sharing for this project was supported by the Washington Heights-Inwood Columbia Aging Project (WHICAP, PO1AG07232, R01AG037212, RF1AG054023) funded by the National Institute on Aging (NIA). This manuscript has been reviewed by WHICAP investigators for scientific content and consistency of data interpretation with previous WHICAP Study publications. We acknowledge the WHICAP study participants and the WHICAP research and support staff for their contributions to this study. This publication was supported by the National Center for Advancing Translational Sciences, National Institutes of Health, through Grant Number UL1TR001873. The content is solely the responsibility of the authors and does not necessarily represent the official views of the NIH. M.A.K. is supported by National Institutes of Health grants: funded by the NIEHS P30ES009089, R01ES030616, and funded by the NIA RF1AG071024 and R01 AG066793. Y.G. is supported by NIA grant R01AG059013.

