## supplemental_material for "Linking air pollution exposure to blood-based metabolic features in a community-based aging cohort"

Supplemental Figures

S Figure 1. Effect modification by Alzheimer's disease

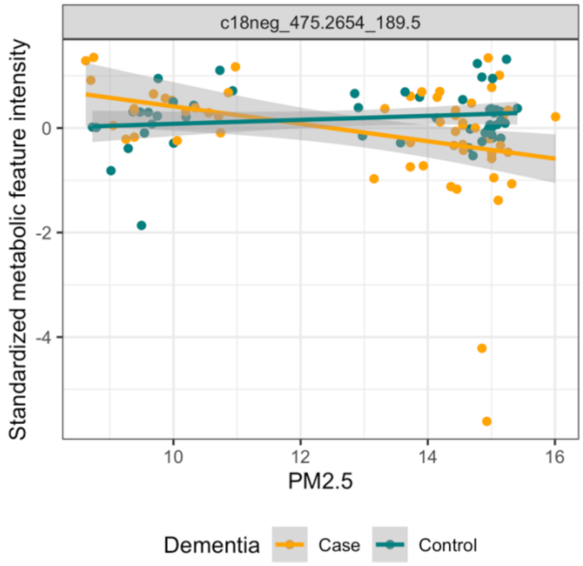

S Figure 2. Effect modification by APOE-ε4 allele status

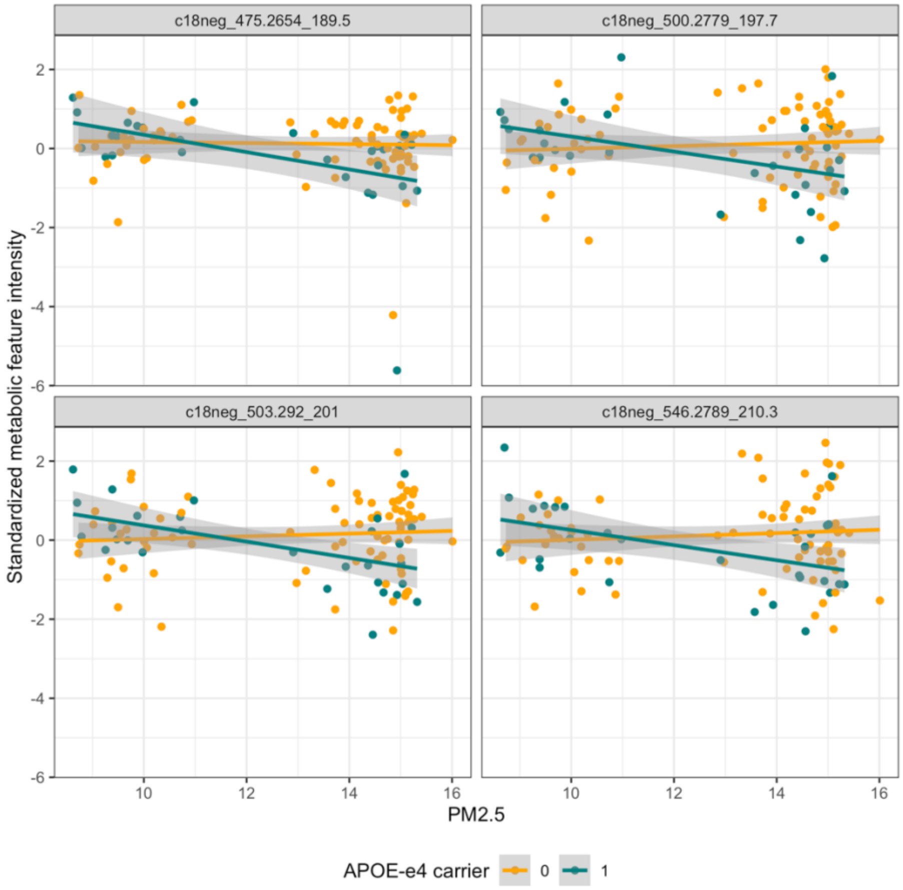

S Figure 3. Effect modification by a history of diabetes

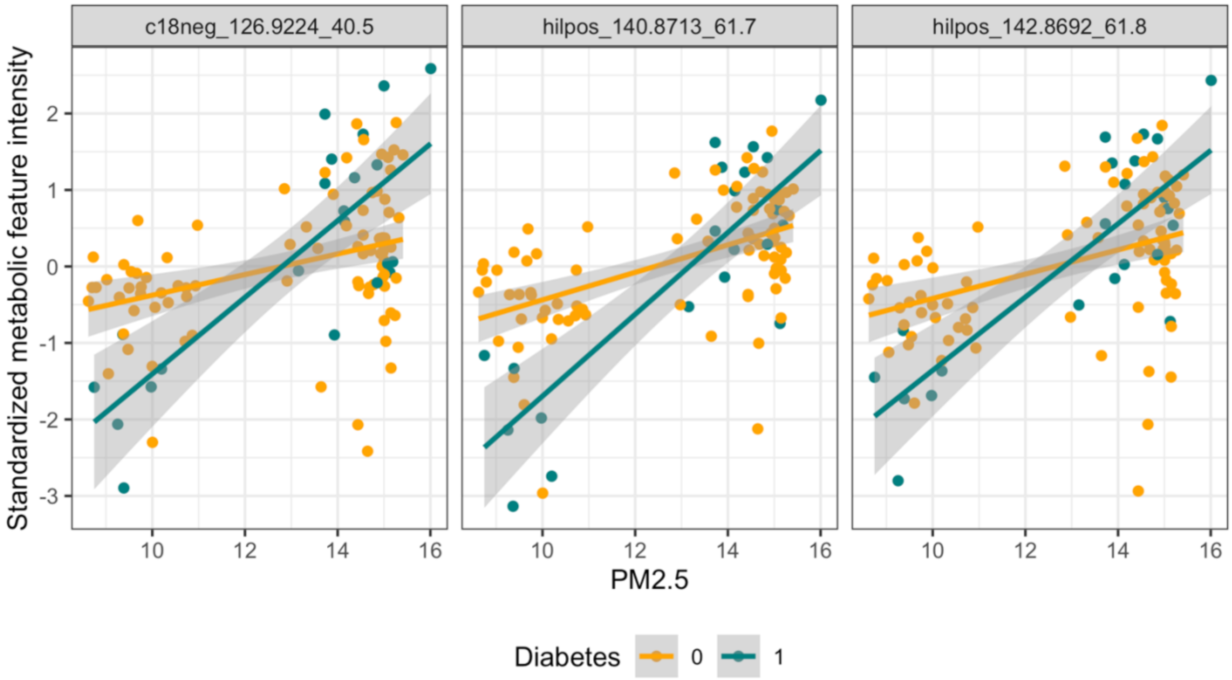

S Figure 4. Principal component analysis of macronutrient dietary data. Vfat: vegetable fat, prot: protein, carbo: carbohydrate, afat: animal fat.

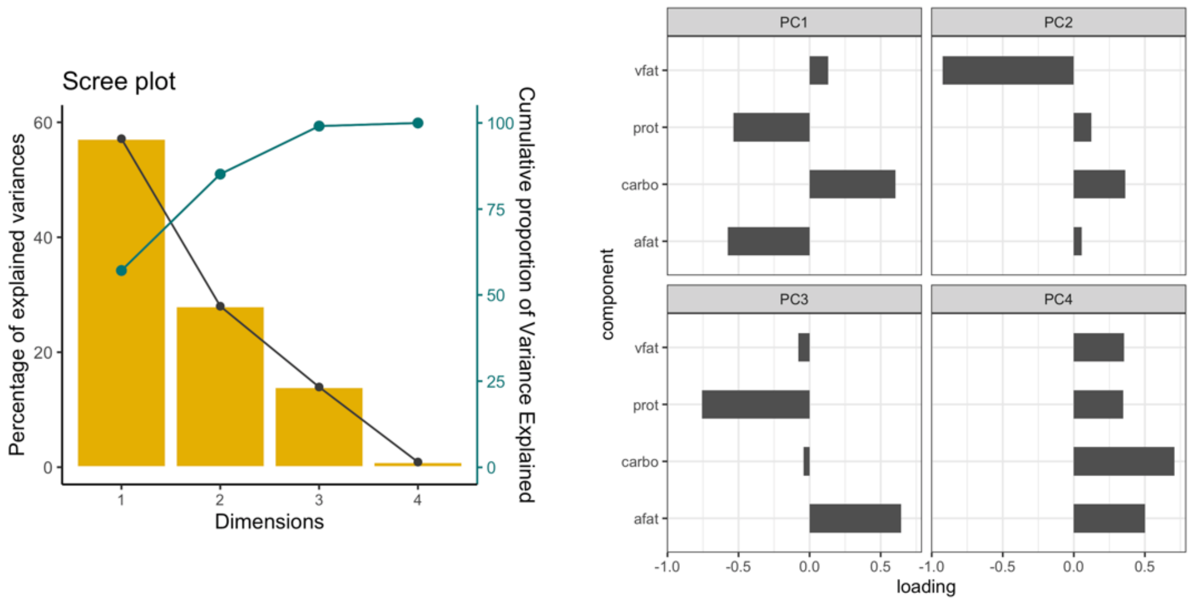

S Figure 5. Sensitivity analysis with smoking history

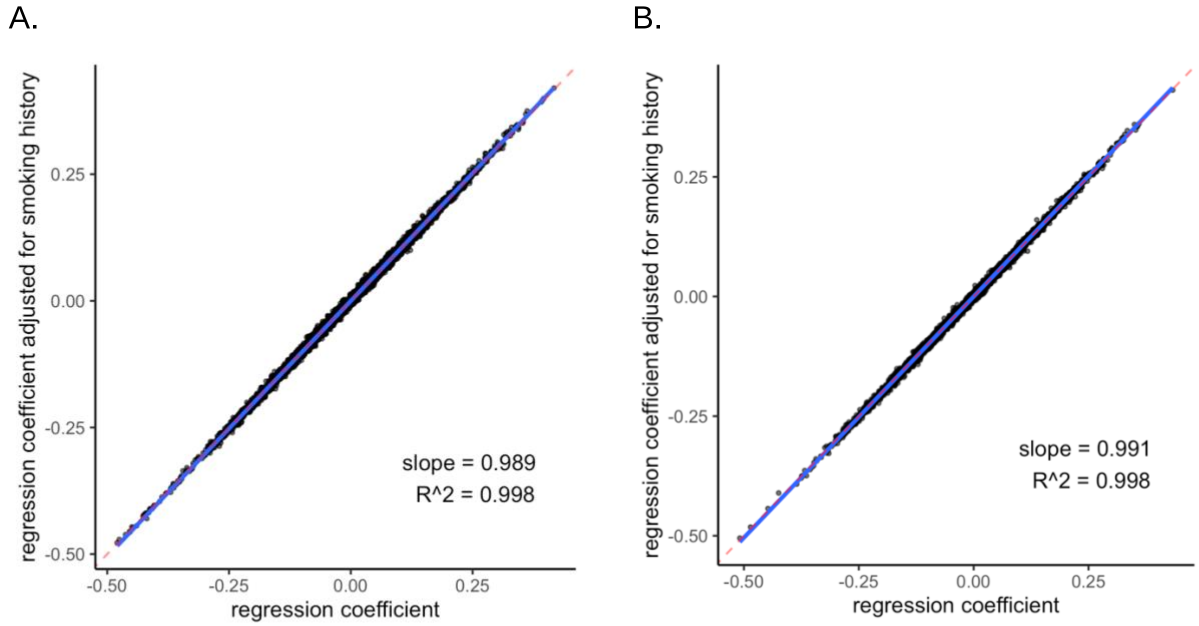


On the x-axis, regression coefficient for $PM_{2.5}$ from model unadjusted for smoking history and on y-axis, regression coefficient for $PM_{2.5}$ from model adjusted for smoking history. A: HILIC + data. B: C18 – data.

Supplemental Tables.

S Table 1. Characteristics of the study population in the Washington Heights and Inwood Community Aging Project comparing cases and controls of Alzheimer's disease.

| | **Case (N=57)** | **Control (N=50)** | **Overall (N=107)** |
|---|---|---|---|
| **Age (years)** | | | |
| Mean (SD) | 86.9 (10.8) | 91.7 (2.93) | 89.2 (8.44) |
| **Sex** | | | |
| Women | 46 (80.7%) | 42 (84.0%) | 88 (82.2%) |
| Men | 11 (19.3%) | 8 (16.0%) | 19 (17.8%) |
| **Racial/Ethnic Group** | | | |
| Non-Hispanic Black | 20 (35.1%) | 17 (34.0%) | 37 (34.6%) |
| Non-Hispanic white | 19 (33.3%) | 17 (34.0%) | 36 (33.6%) |
| Caribbean Hispanic | 18 (31.6%) | 16 (32.0%) | 34 (31.8%) |
| **Education (years)** | | | |
| Mean (SD) | 8.14 (4.66) | 10.9 (4.26) | 9.42 (4.66) |
| **APOE-ε4 carrier** | | | |
| No ε4 allele | 37 (64.9%) | 41 (82.0%) | 78 (72.9%) |
| At least one ε4 allele | 20 (35.1%) | 9 (18.0%) | 29 (27.1%) |
| **History of diabetes** | | | |
| No | 42 (73.7%) | 43 (86.0%) | 85 (79.4%) |
| Yes | 15 (26.3%) | 7 (14.0%) | 22 (20.6%) |
| **History of heart disease** | | | |
| No | 39 (68.4%) | 28 (56.0%) | 67 (62.6%) |
| Yes | 18 (31.6%) | 22 (44.0%) | 40 (37.4%) |
| **History of hypertension** | | | |
| No | 13 (22.8%) | 18 (36.0%) | 31 (29.0%) |
| Yes | 44 (77.2%) | 32 (64.0%) | 76 (71.0%) |
| **$PM_{2.5}$ (μg/m3)** | | | |
| Mean (SD) | 13.0 (2.36) | 12.9 (2.48) | 12.9 (2.41) |
| **PM10 (μg/m3)** | | | |
| Mean (SD) | 20.7 (7.43) | 21.4 (8.12) | 21.0 (7.73) |
| **NO2 (ppb)** | | | |
| Mean (SD) | 31.7 (6.89) | 31.7 (7.25) | 31.7 (7.03) |

S Table 2

Putative annotations of features associated with $PM_{2.5}$ through the metabolome-wide association study framework. m/z: mass-to-charge ratio, Time: Retention time, Delta ppm: mass difference in parts per million, ID score: confidence in annotation based on Schymanski scale (1 being the highest and 5 the lowest), ESI: electrospray ionization.

| m/z | time | ID score | delta ppm | Putative annotation | Adduct | ESI | $\beta$ | q-value |
|---|---|---|---|---|---|---|---|---|
| 243.0263 | 182.7 | 3 | - | Cystine | M+H_[+2] | + | -0.473 | 8.86E-04 |
| 300.0478 | 120.7 | 3 | - | L-Cysteinylglycine disulfide; multiple matches | M+H_[+2] | + | -0.486 | 8.86E-04 |
| 149.53 | 124.3 | 3 | 0.47 | Cysteinylglycine disulfide | M+2H | + | -0.453 | 3.80E-03 |
| 141.5325 | 181.9 | 3 | 0.14 | Cystine | M+ACN+2H | + | -0.427 | 4.29E-03 |
| 298.0517 | 119 | 3 | 2.99 | Cysteinylglycine disulfide | M+H | + | -0.425 | 4.29E-03 |
| 774.5957 | 47.4 | 3 | 6.49 | A phosphatidylcholine or phosphatidylethanolamine | M+H | + | -0.428 | 4.29E-03 |
| 263.0126 | 183.2 | 3 | 1.79 | Cystine | M+Na | + | -0.419 | 5.73E-03 |
| 320.0336 | 122.2 | 3 | 2.91 | L-Cysteinylglycine disulfide | M+Na | + | -0.426 | 5.73E-03 |
| 241.0308 | 181.6 | 3 | 1.33 | Cystine | M+H | + | -0.395 | 8.09E-03 |
| 324.3257 | 41.9 | 3 | 1.2 | N-(14-Methylhexadecanoyl)pyrrolidine | M+H | + | 0.352 | 1.24E-02 |
| 170.0128 | 179.7 | 3 | 6.06 | Cysteic acid | M+H | + | -0.372 | 2.15E-02 |
| 817.5875 | 45.6 | 3 | 2.54 | A phosphatidylethanolamine | M+ACN+H | + | 0.354 | 2.68E-02 |
| 122.027 | 183.9 | 3 | 0.25 | Cysteine | M+H | + | -0.360 | 2.77E-02 |
| 161.0076 | 183.1 | 3 | 8.26 | (2-Furanylmethyl) methyl disulfide | M+H | + | -0.362 | 2.97E-02 |
| 299.0555 | 124.2 | 3 | 1.64 | Aflatoxin P1; Multiple matches | M+H | + | -0.368 | 2.97E-02 |
| 265.0197 | 181.8 | 3 | 4 | 58-Dihydro-6-(4-methyl-3-pentenyl)-1234-tetrathiocin | M+H | + | 0.336 | 3.17E-02 |
| 503.2963 | 49.1 | 3 | - | Fexofenadine; a lysophosphatidylethanolamine | M+H_[+1] | + | -0.358 | 3.80E-02 |

| | | | | | | | | |
|---|---|---|---|---|---|---|---|---|
| 309.2056 | 40.5 | 3 | 1.42 | Multiple matches | M+H | + | 0.276 | 4.77E-02 |
| 190.0357 | 35.6 | 3 | 0.05 | Isosorbide Mononitrate; gamma-Carboxyglutamic acid | M-H | - | -0.506 | 4.71E-04 |
| 129.0194 | 36.5 | 3 | 0.54 | A dicarboxylic acid | M-H | - | -0.488 | 9.22E-04 |
| 177.0041 | 35.4 | 3 | 4.75 | Bissulfine; multiple other matches | | - | -0.431 | 4.60E-03 |
| 209.0303 | 35.4 | 3 | 0.05 | A dicarboxylic acid | M-H | - | -0.446 | 5.22E-03 |
| 475.2654 | 189.5 | 3 | - | A lysophosphatidylethanolamine | M-H_[-1] | - | -0.353 | 1.66E-02 |
| 503.292 | 201 | 3 | - | A lysophosphatidylethanolamine | M-H_[-1] | - | -0.381 | 1.83E-02 |
| 500.2779 | 197.7 | 3 | 0.72 | A lysophosphatidylethanolamine | M-H | - | -0.383 | 2.71E-02 |
| 502.2937 | 205 | 3 | 0.42 | A lysophosphatidylethanolamine | M-H | - | -0.341 | 4.94E-02 |
| 154.9871 | 183.3 | 5 | 8 | Alphachloralose | M+2H | + | -0.454 | 1.46E-03 |
| 355.0733 | 115.4 | 5 | 1.75 | norsertraline; multiple matches | M+ACN+Na | + | -0.444 | 3.86E-03 |
| 361.048 | 121.3 | 5 | 8.5 | 6-Demethylgriseofulvin | M+Na | + | -0.424 | 4.18E-03 |
| 775.6017 | 48 | 5 | 4.32 | A diglyceride | M+2ACN+H | + | -0.429 | 4.18E-03 |
| 98.5181 | 183.4 | 5 | 3.25 | Diethyl trisulfide, multiple other matches | M+ACN+2H | + | -0.419 | 5.73E-03 |
| 152.9913 | 182.8 | 5 | 1.24 | Roxarsone | M+ACN+2H | + | -0.413 | 5.73E-03 |
| 360.0233 | 98.5 | 5 | 0.47 | Cefdinir | M+H-2H2O | + | -0.406 | 5.73E-03 |
| 822.5468 | 54.6 | 5 | 3.65 | A phosphatidylethanolamine | M+H-H2O | + | 0.400 | 5.85E-03 |
| 178.0407 | 118.6 | 5 | 1.35 | Methimazole | M+ACN+Na | + | -0.417 | 6.23E-03 |
| 824.5603 | 55.7 | 5 | 1.02 | A phosphatidylethanolamine | M+H-H2O | + | 0.399 | 9.80E-03 |
| 296.2944 | 42.1 | 5 | | Multiple matches | | + | 0.319 | 1.10E-02 |
| 132.9872 | 60.1 | 5 | 0 | Pyruvic acid, multiple other matches | M+2Na-H | + | -0.395 | 1.11E-02 |
| 195.0259 | 184.9 | 5 | 1.28 | Thiocysteine | M+ACN+H | + | -0.388 | 1.18E-02 |
| 130.159 | 43.8 | 5 | 0.23 | Decamethonium/Propane | M+2H | + | 0.362 | 1.58E-02 |
| 378.7789 | 61.9 | 5 | 7.42 | A diglyceride | M+ACN+2H | + | 0.332 | 2.77E-02 |
| 586.0316 | 182.9 | 5 | 8.75 | Zoledronate | 2M+ACN+H | + | -0.371 | 2.77E-02 |
| 98.0164 | 186.8 | 5 | 0.61 | Thiocysteine | M+ACN+2H | + | -0.357 | 2.97E-02 |
| 142.0342 | 182.5 | 5 | 2.53 | Polyvidone | M+H-2H2O | + | -0.362 | 2.97E-02 |

| | | | | | | | | |
|---|---|---|---|---|---|---|---|---|
| 461.3484 | 48 | 5 | 3.97 | Solanidine | M+ACN+Na | + | 0.307 | 2.97E-02 |
| 242.0344 | 181.1 | 5 | 3.76 | Glucosamine 6-sulfate | M+H-H2O | + | -0.359 | 3.70E-02 |
| 130.5333 | 26.1 | 5 | 2.3 | Tyrosol 4-sulfate | M+ACN+2H | + | -0.161 | 4.26E-02 |
| 796.5254 | 57.2 | 5 | 0.3 | A phosphatidylethanolamine | M+Na | + | 0.344 | 4.26E-02 |
| 244.2632 | 42.7 | 5 | 1.19 | Pentadecanal; 2-Pentadecanone; Heptyl ketone | M+NH4 | + | 0.343 | 4.86E-02 |
| 264.0158 | 183.5 | 5 | 5.49 | 5-Sulfo-13-benzenedicarboxylic acid | M+NH4 | + | -0.357 | 4.86E-02 |
| 267.0003 | 36.7 | 5 | 0.94 | Cyclobrassinone | M+Cl | - | 0.367 | 2.39E-02 |
| 605.3636 | 229.7 | 5 | 3.57 | A Hericenone | M+Cl | - | -0.390 | 2.39E-02 |
| 265.0024 | 37.2 | 5 | 1.96 | Apraclonidine | M+Na-2H | - | 0.360 | 2.55E-02 |
| 356.9921 | 31.7 | 5 | 4.82 | Cyanidin | M+Cl | - | 0.352 | 4.76E-02 |
| 320.0038 | 182.8 | | | *Unknown* | | + | -0.481 | 8.86E-04 |
| 120.0115 | 183.7 | | | *Unknown* | | + | -0.450 | 1.46E-03 |
| 162.5474 | 182.9 | | | *Unknown* | | + | -0.460 | 1.68E-03 |
| 121.0192 | 186.1 | | | *Unknown* | | + | -0.433 | 4.18E-03 |
| 140.8713 | 61.7 | | | *Unknown* | | + | 0.359 | 4.29E-03 |
| 162.0457 | 185.1 | | | *Unknown* | | + | -0.421 | 4.29E-03 |
| 288.2889 | 42.7 | | | *Unknown* | | + | 0.419 | 8.09E-03 |
| 587.035 | 182.7 | | | *Unknown* | | + | -0.406 | 8.83E-03 |
| 156.8449 | 62 | | | *Unknown* | | + | 0.391 | 9.69E-03 |
| 185.5406 | 273.4 | | | *Unknown* | | + | -0.382 | 1.38E-02 |
| 142.5304 | 184.8 | | | *Unknown* | | + | -0.372 | 1.58E-02 |
| 181.0193 | 183.2 | | | *Unknown* | | + | -0.378 | 1.58E-02 |
| 142.8692 | 61.8 | | | *Unknown* | | + | 0.344 | 1.74E-02 |
| 86.1112 | 57.6 | | | *Unknown* | | + | 0.342 | 2.44E-02 |
| 160.506 | 183.2 | | | *Unknown* | | + | -0.357 | 2.77E-02 |
| 121.5209 | 183.1 | | | *Unknown* | | + | -0.362 | 3.10E-02 |
| 852.6074 | 57.8 | | | *Unknown* | | + | 0.340 | 4.74E-02 |

| | | | | | |
|---|---|---|---|---|---|
| 872.5548 | 57.2 | *Unknown* | + | 0.359 | 4.74E-02 |
| 866.5841 | 60.4 | *Unknown* | + | 0.328 | 4.86E-02 |
| 109.0224 | 25.8 | *Unknown* | + | -0.335 | 5.03E-02 |
| 126.9224 | 40.5 | *Unknown* | - | 0.437 | 1.61E-03 |
| 168.836 | 35.3 | *Unknown* | - | 0.353 | 5.22E-03 |
| 479.2843 | 183.6 | *Unknown* | - | -0.367 | 1.87E-02 |
| 280.8711 | 42.1 | *Unknown* | - | 0.364 | 2.39E-02 |
| 546.2789 | 210.3 | *Unknown* | - | -0.382 | 2.61E-02 |
| 500.4841 | 197.4 | *Unknown* | - | -0.372 | 4.94E-02 |

S Table 3

Putative annotations of features with a non-zero coefficient when predicting $PM_{2.5}$ exposure through the elastic net regression. m/z: mass-to-charge ratio, Time: Retention time, Delta ppm: mass difference in parts per million, ID score: confidence in annotation based on Schymanski scale (1 being the highest and 5 the lowest), ESI: electrospray ionization.

| m/z | time | ID score | delta ppm | Putative annotation | Adduct | ESI | $\beta$ | q-value |
|---|---|---|---|---|---|---|---|---|
| 190.0357 | 35.6 | 3 | 0.05 | Isosorbide Mononitrate; gamma-Carboxyglutamic acid | M-H | - | -0.51 | 0.0003 |
| 129.0194 | 36.5 | 3 | 0.54 | A dicaroxylic acid | M-H | - | -0.49 | 0.0009 |
| 354.9404 | 33.3 | 3 | 5.75 | Silver sulfadiazine | M-H | - | 0.22 | 0.2751 |
| 387.33 | 290.9 | 3 | 8.13 | 3-Hydroxy-1-phenyl-1-eicosanone | M-H | - | 0.28 | 0.1453 |
| 149.53 | 124.3 | 3 | 0.47 | L-Cysteinylglycine disulfide | M+2H | + | -0.45 | 0.0039 |
| 300.0478 | 120.7 | 3 | - | L-Cysteinylglycine disulfide; Glyzaglabrin; Aflatoxin P1 | M+H_[+2] | + | -0.48 | 0.0011 |
| 167.0825 | 60.9 | 3 | - | L-Phenylalanine; multiple other matches | M+H_[+1] | + | -0.29 | 0.1533 |
| 163.1329 | 22.7 | 3 | 0.18 | (3R7R)-137-Octanetriol | M+H | + | -0.21 | 0.4118 |
| 508.3385 | 53.8 | 3 | 2.5 | A lysophosphatidylethanolamine | M+H | + | -0.23 | 0.3093 |
| 138.0914 | 228.7 | 3 | 0.43 | Tyramine; multiple other matches | M+H | + | -0.28 | 0.2326 |
| 205.0171 | 25.9 | 3 | 2.83 | O-methoxycatechol-O-sulphate | M+H | + | -0.34 | 0.0494 |
| 605.3636 | 229.7 | 5 | 3.57 | A Hericenone | M+Cl | - | -0.39 | 0.0309 |
| 98.0964 | 56.2 | 5 | 0.31 | Ethylene; 3-Nonyl-1H-pyrazole | 2M+ACN+H; M+2H | + | -0.27 | 0.1519 |
| 130.159 | 43.8 | 5 | 0.23 | Decamethonium; Propane | M+2H; 2M+ACN+H | + | 0.35 | 0.0228 |
| 154.9871 | 183.3 | 5 | 8 | Alphachloralose | M+2H | + | -0.45 | 0.0015 |
| 355.0733 | 115.4 | 5 | 1.75 | norsertraline | M+ACN+Na | + | -0.44 | 0.0043 |
| 556.4413 | 40.5 | 5 | 3.4 | Didodecyl thiobispropanoate | M+ACN+H | + | 0.28 | 0.2031 |
| 775.6017 | 48 | 5 | 4.32 | a Diacylglyceride | M+2ACN+H | + | -0.42 | 0.0057 |
| 114.9341 | 144.4 | - | - | *Unknown* | | - | -0.29 | 0.1716 |
| 126.9224 | 40.5 | - | - | *Unknown* | | - | 0.43 | 0.0018 |
| 137.8919 | 112.2 | - | - | *Unknown* | | - | -0.28 | 0.0632 |

| | | | | | | | |
|---|---|---|---|---|---|---|---|
| 236.9005 | 85.7 | - | - | *Unknown* | - | -0.28 | 0.1432 |
| 254.9444 | 45.3 | - | - | *Unknown* | - | -0.36 | 0.0512 |
| 279.6142 | 34.1 | - | - | *Unknown* | - | -0.31 | 0.0956 |
| 479.2843 | 183.6 | - | - | *Unknown* | - | -0.37 | 0.0183 |
| 500.4841 | 197.4 | - | - | *Unknown* | - | -0.37 | 0.0507 |
| 796.5517 | 220.4 | - | - | *Unknown* | - | 0.35 | 0.0837 |
| 546.2789 | 210.3 | - | - | *Unknown* | - | -0.38 | 0.0261 |
| 179.9855 | 23.6 | - | - | *Unknown* | + | -0.29 | 0.1751 |
| 288.2889 | 42.7 | - | - | *Unknown* | + | 0.42 | 0.0083 |
| 470.9033 | 73 | - | - | *Unknown* | + | 0.31 | 0.0757 |
| 646.7869 | 70.4 | - | - | *Unknown* | + | -0.32 | 0.0971 |
| 202.9893 | 288.2 | | | *Unknown* | + | 0.31 | 0.1143 |

S Table 4

Putative annotations of features with high variable importance scores (VIP) in predicting high or low $PM_{2.5}$ exposure groups through the partial least square discriminant analysis. m/z: mass-to-charge ratio, Time: Retention time, Delta ppm: mass difference in parts per million, ID score: confidence in annotation based on Schymanski scale (1 being the highest and 5 the lowest), ESI: electrospray ionization. VIP comp1: variable importance score on component 1, VIP comp2: variable importance score on component 1

| mz | time | ID score | delta ppm | Putative annotation | Adduct | ESI | $\beta$ | q-value | VIP comp1 | VIP comp2 |
|---|---|---|---|---|---|---|---|---|---|---|
| 300.0478 | 120.7 | 3 | - | L-Cysteinylglycine disulfide; multiple matches | M+H_[+2] | + | -0.486 | 8.86E-04 | 4.18 | 3.16 |
| 243.0263 | 182.7 | 3 | - | Cystine | M+H_[+2] | + | -0.473 | 8.86E-04 | 4.14 | 3.02 |
| 177.0041 | 35.4 | 3 | 4.75 | Bissulfine; multiple other matches | | - | -0.431 | 4.60E-03 | 3.93 | 2.96 |
| 129.0194 | 36.5 | 3 | 0.54 | A dicarboxylic acid | M-H | - | -0.488 | 9.22E-04 | 3.71 | 2.72 |
| 149.53 | 124.3 | 3 | 0.47 | Cysteinylglycine disulfide | M+2H | + | -0.453 | 3.80E-03 | 3.70 | 2.77 |
| 320.0336 | 122.2 | 3 | 2.91 | L-Cysteinylglycine disulfide | M+Na | + | -0.426 | 5.73E-03 | 3.44 | 2.59 |
| 263.0126 | 183.2 | 3 | 1.79 | Cystine | M+Na | + | -0.419 | 5.73E-03 | 3.36 | 2.41 |
| 276.1114 | 44.2 | 3 | 4.31 | Queuine | M-H | - | 0.266 | 1.28E-01 | 3.28 | 2.68 |
| 241.0308 | 181.6 | 3 | 1.33 | Cystine | M+H | + | -0.395 | 8.09E-03 | 3.27 | 2.38 |
| 298.0517 | 119 | 3 | 2.99 | Cysteinylglycine disulfide | M+H | + | -0.425 | 4.29E-03 | 3.22 | 2.43 |
| 209.0303 | 35.4 | 3 | 0.05 | A dicarboxylic acid | M-H | - | -0.446 | 5.22E-03 | 3.10 | 2.25 |
| 214.0513 | 253.3 | 3 | 0.33 | Cysteineglutathione disulfide | M+2H | + | -0.335 | 6.47E-02 | 3.09 | 2.34 |
| 141.5325 | 181.9 | 3 | 0.14 | Cystine | M+ACN+2H | + | -0.427 | 4.29E-03 | 3.06 | 2.19 |
| 299.0555 | 124.2 | 3 | 1.64 | Aflatoxin P1; Multiple matches | M+H | + | -0.368 | 2.97E-02 | 3.02 | 2.24 |
| 301.0511 | 119.9 | 3 | - | L-Cysteinylglycine disulfide; multiple other matches | M+H_[+3] | + | -0.342 | 5.99E-02 | 3.00 | 2.36 |
| 236.0894 | 286.7 | 3 | 9.87 | 3-Oxo-carbofuran | M+H | + | 0.133 | 6.18E-01 | 2.97 | 2.51 |
| 355.0733 | 115.4 | 5 | 1.75 | norsertraline; multiple matches | M+ACN+Na | + | -0.444 | 3.86E-03 | 4.21 | 3.30 |
| 361.048 | 121.3 | 5 | 8.5 | 6-Demethylgriseofulvin | M+Na | + | -0.424 | 4.18E-03 | 4.00 | 3.00 |
| 178.0407 | 118.6 | 5 | 1.35 | Methimazole | M+ACN+Na | + | -0.417 | 6.23E-03 | 3.96 | 3.20 |

| | | | | | | | | | | |
|---|---|---|---|---|---|---|---|---|---|---|
| 195.0259 | 184.9 | 5 | 1.28 | Thiocysteine | M+ACN+H | + | -0.388 | 1.18E-02 | 3.51 | 2.56 |
| 775.6017 | 48 | 5 | 4.32 | A diglyceride | M+2ACN+H | + | -0.429 | 4.18E-03 | 3.35 | 2.54 |
| 154.9871 | 183.3 | 5 | 8 | Alphachloralose | M+2H | + | -0.454 | 1.46E-03 | 3.26 | 2.34 |
| 98.0164 | 186.8 | 5 | 0.61 | Thiocysteine | M+ACN+2H | + | -0.357 | 2.97E-02 | 3.18 | 2.29 |
| 98.5181 | 183.4 | 5 | 3.25 | Diethyl trisulfide, multiple other matches | M+ACN+2H | + | -0.419 | 5.73E-03 | 3.12 | 2.23 |
| 152.9913 | 182.8 | 5 | 1.24 | Roxarsone | M+ACN+2H | + | -0.413 | 5.73E-03 | 3.08 | 2.22 |
| 264.0158 | 183.5 | 5 | 5.49 | 5-Sulfo-13-benzenedicarboxylic acid | M+NH4 | + | -0.357 | 4.86E-02 | 3.05 | 2.18 |
| 826.685 | 71.8 | 5 | 3.5 | N-Glycoloylganglioside GM2 | M+2ACN+H | + | -0.248 | 2.91E-01 | 3.03 | 2.35 |
| 120.0115 | 183.7 | | | *Unknown* | | + | -0.450 | 1.46E-03 | 4.17 | 3.06 |
| 320.0038 | 182.8 | | | *Unknown* | | + | -0.481 | 8.86E-04 | 4.16 | 3.04 |
| 162.0457 | 185.1 | | | *Unknown* | | + | -0.421 | 4.29E-03 | 3.71 | 2.67 |
| 181.0193 | 183.2 | | | *Unknown* | | + | -0.378 | 1.58E-02 | 3.41 | 2.45 |
| 162.5474 | 182.9 | | | *Unknown* | | + | -0.460 | 1.68E-03 | 3.28 | 2.36 |
| 121.0192 | 186.1 | | | *Unknown* | | + | -0.433 | 4.18E-03 | 3.27 | 2.35 |
| 160.506 | 183.2 | | | *Unknown* | | + | -0.357 | 2.77E-02 | 3.20 | 2.29 |
| 185.5406 | 273.4 | | | *Unknown* | | + | -0.382 | 1.38E-02 | 3.13 | 2.24 |
| 181.5296 | 121.4 | | | *Unknown* | | + | -0.309 | 6.94E-02 | 3.11 | 2.38 |
